# Long-term Complications of Unintentional Dural Puncture During Labor Epidural Analgesia: A Case-Control Study

**DOI:** 10.1101/2021.10.18.21265155

**Authors:** Alexandre Lacombe, Kristi Downey, Xiang Y. Ye, Jose C.A. Carvalho

## Abstract

**Introduction:** Epidural analgesia is the preferred method to manage pain during labor and delivery. The insertion of the epidural catheter can be complicated by unintentional dural puncture that may result in postdural puncture headache. There is limited evidence on the long term implications of this complication. We sought to investigate if women who sustained a dural puncture have a higher risk of developing chronic headache, low back pain and visual or auditory impairment.

**Methods:** We conducted a 1:1 case-control study with women who delivered at our institution from January 2015 to December 2019. Cases were women who received epidural analgesia and sustained an unintentional dural puncture and controls were women who received epidural analgesia but did not sustain such complication. We matched cases and controls for date of delivery, age, and BMI. All women completed an online survey with validated questionnaires for diagnosis of chronic headache and chronic back pain. We used dichotomic (yes/no) questions to look for the presence of chronic visual and auditory impairment.

**Results:** Sixty-three case-control pairs were studied. Women who sustained a dural puncture during their epidural catheter insertion had a higher risk of developing chronic headache [14.3%, versus 4.8%, p=0.049, AOR: 3.36 (1.05, 12.82)] and chronic back pain [39.7% versus 19.1%, p=0.009, AOR: 2.67 (1.25, 5.72)] than women who did not sustain a dural puncture. The incidence of chronic auditory impairment was also higher in the dural puncture group [14.3% versus 1.6%, p=0.007, AOR: 9.98 (1.21, 82.62)].

**Conclusions:** An unintentional dural puncture during epidural catheter insertion in parturients is associated with increased risk of chronic headache, back pain and auditory impairment.

## Introduction

Epidural analgesia is commonly used to manage pain in childbirth. The technique is efficient, safe and allows conversion to surgical anesthesia if necessary. At our institution, the rate of epidural analgesia in laboring women was 88% between July 2019 and June 2020.

The epidural catheter insertion may be associated with complications. Among those, an unintentional dural puncture (UDP) may occur at a rate of 0.5% to 1.5% of all insertions ^(1-2)^. This complication is frequently associated with postdural puncture headache (PDPH), a positional and sometimes incapacitating headache that may be accompanied by nausea, vomiting, neck stiffness and visual and/or auditory disturbances ^(3)^. The symptoms may be self-limited and managed by conservative measures. If the latter fail, the epidural blood patch (EBP) has shown benefit ^(4)^ and is regarded by many as the ultimate treatment for PDPH.

The symptoms associated with UDP will resolve completely in most women, but some might develop long-term complications. Indeed, PDPH has been associated with chronic headache and chronic lower backache ^(5-7)^. Furthermore, the effect of EBP on chronic low back pain and headache in women who sustained UDP is still controversial ^(5-6)^.

Despite the widespread use of epidural analgesia for labor pain management and the frequency of UDP, there is a paucity of data on long-term complications associated with UDP and EBP. We sought to investigate whether women sustaining UDP at our institution developed long-term complications. Furthermore, we sought to investigate the effect of EBP on their outcomes.

Our hypothesis was that women who sustained UDP during labor epidural catheter insertion would have a higher risk of developing long-term complications when compared with those who received epidural analgesia but did not sustain UDP.

## Methods

This 1:1 case-control study was conducted with approval of the Ethics Review Board at Mount Sinai Hospital in Toronto, Ontario, Canada (REB#20-0278-E; January 20^th^, 2021) and patient informed consent. We studied women who delivered at our institution from January 2015 to December 2019 and requested epidural analgesia for labor pain management and either sustained UDP (cases) or not (controls).

Management of UDP: Since this was a retrospective study, the management of UDP varied at the discretion of the attending anesthesiologist. At our institution women who sustain UDP and develop PDPH are offered a conservative management including acetaminophen, non-steroidal anti-inflammatories, in addition to opioids if necessary. Some anesthesiologists may proceed with the insertion of an intrathecal catheter, especially if the epidural insertion was challenging. Patients are followed by daily visits while in hospital, or by daily phone calls after discharge. They are offered a therapeutic epidural blood patch after approximately 48hrs after the UDP if the severity of the symptoms prevents them from performing their daily activities (nursing the newborn for example) or if there are visual or auditory symptoms. The follow-up continues until improvement or resolution of symptoms. All women who sustain an UDP are discharged with an information document including the typical characteristics of PDPH, a reminder of the conservative measures and a phone number to call if the headache persists or worsens after their hospital discharge. They are also instructed to call should they develop any visual or auditory impairment after discharge.

The cases were identified from a database of women sustaining or suspected of having sustained UDP, maintained by the Department of Anesthesia at Mount Sinai Hospital. We only included cases of recognized UDP, defined as the visualization of cerebral spinal fluid through the 17-gauge Tuohy needle or through the epidural catheter at the moment of the epidural catheter insertion. We excluded women with a history of pre-existing chronic headache or backache. We reviewed their medical records to certify that they met the inclusion criteria, and that the exclusion criteria were not applicable. We looked at demographic (age, height, weight, BMI, gravida and para status) and clinical data of interest (mode of delivery, use of spine ultrasound, number of attempts during epidural catheter insertion and presence of scoliosis) for our study. Once these women were identified, they were approached by phone to explain the study and to ask for consent. Those who could be reached by phone and who consented to participate in the study were requested to complete an online survey for the outcomes of interest.

Once case women were recruited, we matched them 1-1 to a control group of women who received epidural analgesia during labor but did not sustain UDP. Cases and controls were matched by date of delivery (same month, same year), age (same year of birth) and BMI (< 40 or ≥ 40). For the latter, we sometimes had to match women with BMI ≥ 40 to a control with a different month of delivery and a different age (up to ten years difference) because we did not have a patient with these same values in our database. Similar to those in the case group, women in the control group had their medical records reviewed to check for inclusion and exclusion criteria, after which they were approached by phone to explain the study and ask for consent. Those consenting were requested to complete the same online survey.

The cases and their matched controls were recruited at least six months after the epidural placement, to ensure that chronicity of complications was well established.

The online survey was created using Simple Survey (Outsidesoft Solutions inc., Quebec, Canada). The survey can be seen in appendix.

The questions included in the survey were derived from validated questionnaires ^(5, 8-15)^ for the majority of our outcomes of interest. For the few outcomes that a validated questionnaire did not exist we looked for the presence of our endpoint by asking dichotomic (yes/no) questions. We tried to maintain the objectivity of the patient’s answer by asking if the outcomes were self-diagnosed or diagnosed by a healthcare professional.

The primary outcome of our study was the presence of chronic headache defined as headache that lasted or recurred for more than three months. Secondary outcomes were the presence of chronic lower back pain (with or without neuropathic features), chronic auditory or visual impairment and severity of the long-term disability caused by chronic pain Chronicity was defined as symptom that lasted or recurred for more than three months. We also looked at a composite outcome, which was defined as the presence of any of the chronic symptoms including headache, back pain, auditory or visual impairment.

To assess the primary outcome, we used a questionnaire derived from the Chronic Pain Grade Questionnaire, designed to measure chronic pain for severity, persistence, and disability ^(9)^

We used different questionnaires for the secondary outcomes. To evaluate back pain, we used questions from the Chronic Pain Grade Questionnaire ^(9)^ and also the Low Back Pain Rating Scale ^(8)^. To establish if patients developed neuropathic pain from their backache disorders, we used questions from the “Douleur Neuropathique 4 (DN4)” questionnaire ^(14)^. The DN4 questionnaire is one of the most widely used and reliable screening questionnaires and is reported to have good diagnostic properties ^(14)^. We did not have validated questionnaires for auditory and visual symptoms associated with PDPH, so we proceeded with dichotomic “yes” or “no” questions to indicate the presence of these outcomes. We also identified whether the outcome of interest was self-reported or diagnosed by a healthcare professional.

Finally, we used questions derived from the Pain Disability Index (PDI) Questionnaire ^(15)^ to evaluate the impact of the most bothersome symptoms in the patient’s daily activities. The PDI Questionnaire assesses the disability in 7 different aspects of daily activities for a total score of 70.

### Sample size calculation and Statistical Analysis

Since we had no previous information available for sample size estimation for our primary outcome, we reviewed all UDP cases in our database from January 2015 to December 2019. Those satisfying inclusion and exclusion criteria and consenting to participate in the study were included in our case group. We then matched the cases with equal number of controls.

The study population was summarized descriptively. The patient characteristics were compared between the case and control groups using Chi-square test for categorical variables and student t test or Wilcoxon rank sum test for continuous variables as appropriate. To examine the differences in the binary outcomes between the two groups, they were compared between two groups using linear probability model with generalized estimated equation (GEE) approach accounting for the correlation between matched case-control. We also conducted multivariable conditional logistic regression analysis to further compare the binary outcomes adjusted for potential confounders identified in the univariate analyses. For comparing the difference in the continuous outcome of the total PDI score, due to highly skew, quantile regression model without/with adjustment for the potential confounders were applied. Subgroup analyses were also conducted to compare the outcomes including side effects between UDP patients with/without EBP. Similar methods were used for the comparisons. The data management and all statistical analyses were performed using SAS 9.4 (SAS Institute Inc., Cary, NC, USA). A two-sided p-value of <0.05 was used to determine the statistical significance.

## Results

We identified 162 women who sustained UDP from January 2015 to December 2019. Amongst them, 60 could not be reached, 14 declined to participate and 2 were ineligible (language barrier). Eighty-six women consented to participate and 82 completed the survey but 19 were excluded based on their answers (pre-existing backache or headache). Finally, 63 women were included for analysis in the case group. For the control group, we initially contacted 209 women but 112 were excluded because they refused, were not reachable or were ineligible to participate. From the 97 women who consented, 94 completed the survey but 31 were ineligible based on their response (pre-existing backache or headache). A total of 63 women were included in the control group for analysis. The description of patient recruitment can be seen in figure 1.

**Figure 1:**
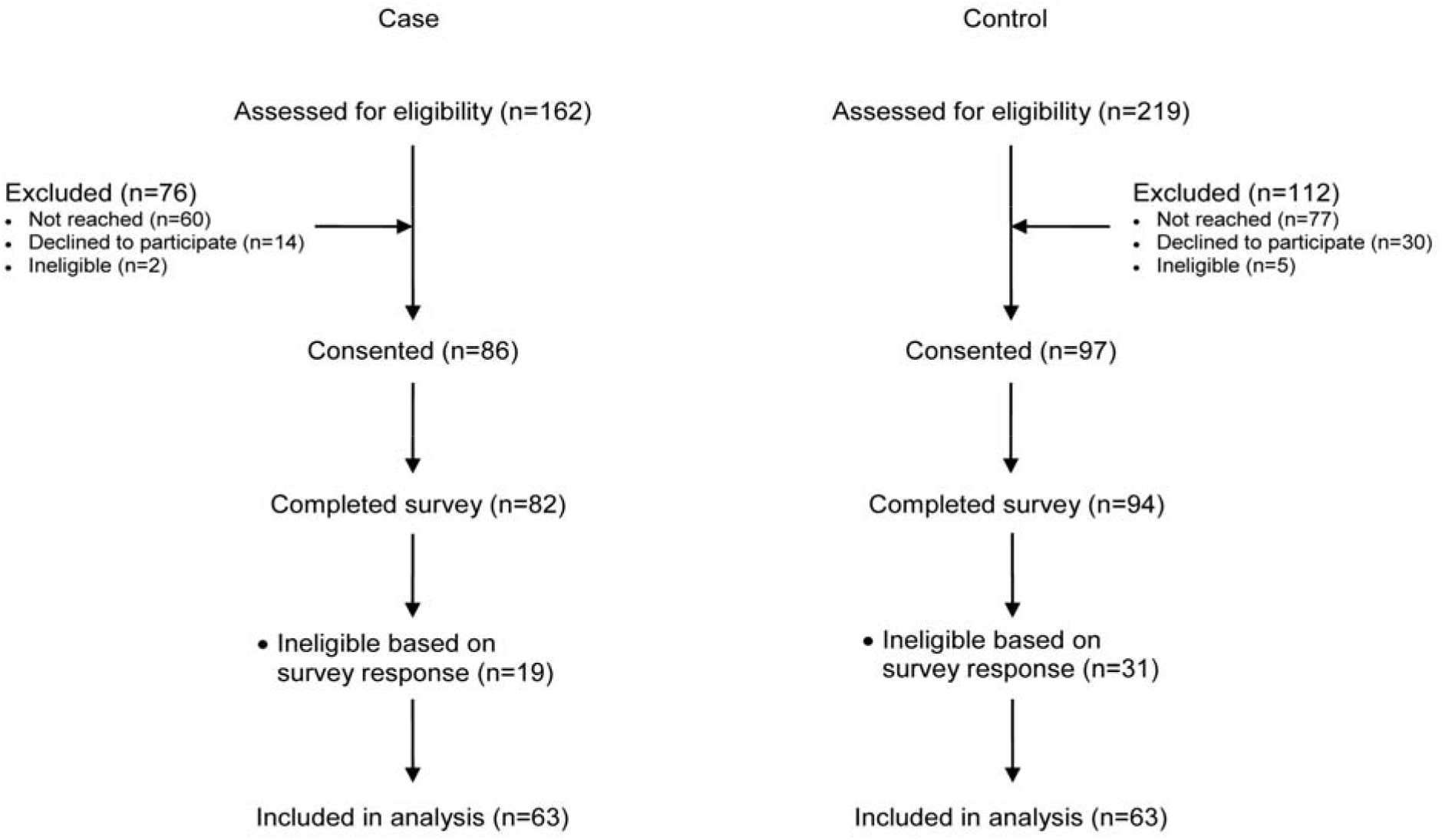
Study Flowchart

There was no difference in age, height and weight, BMI, mode of delivery, presence of scoliosis and use of ultrasound for epidural insertion assistance between the dural puncture group and the control group (Table 1). The number of documented attempts needed for the epidural insertion and the occurrence of paresthesia were more frequent in the UDP group. We identified only one intravascular catheter and it was in the control group. The time between the unintentional dural puncture and the survey completion by the patient varied from 1-6 years.

**Table 1.**
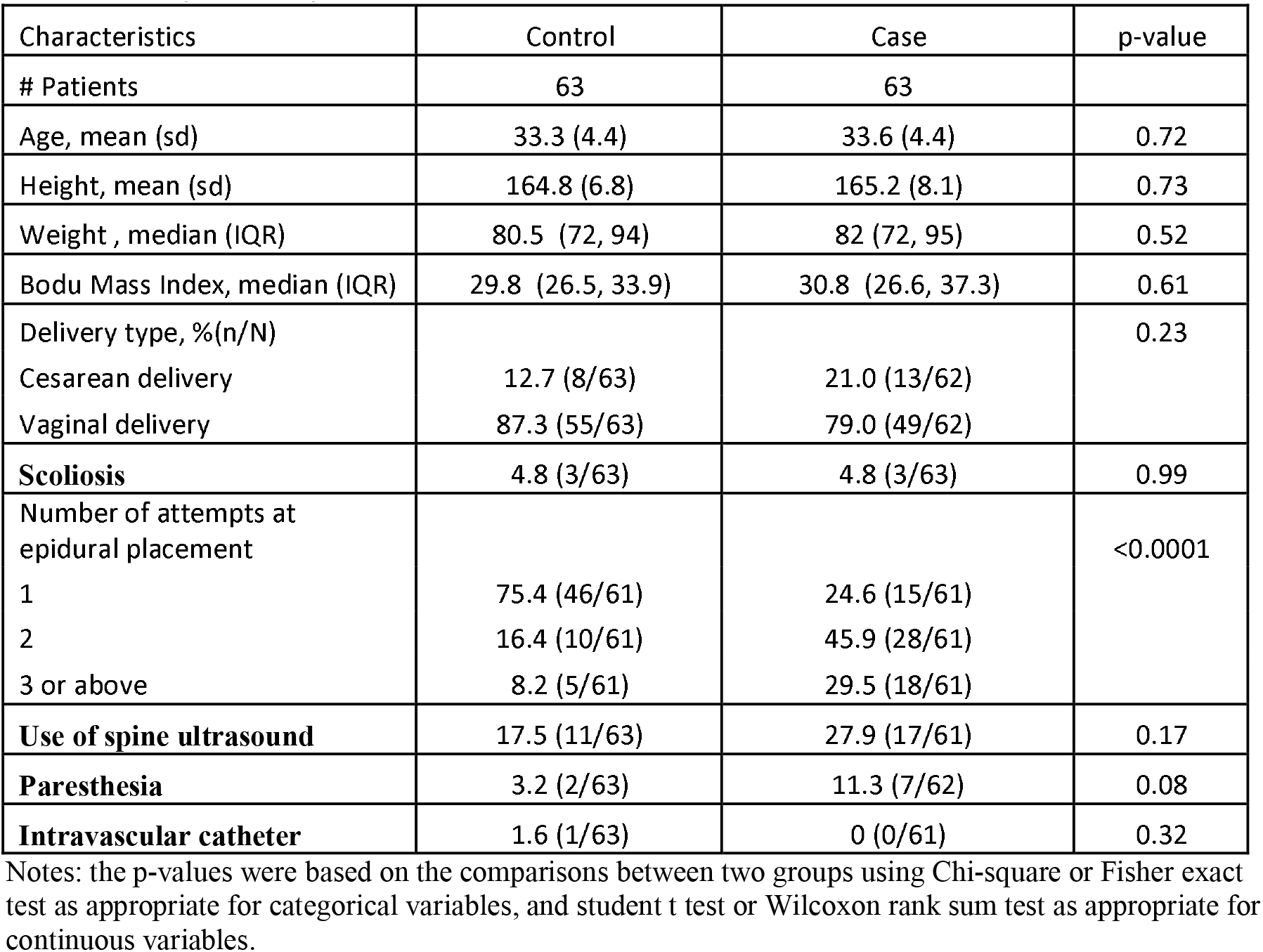
Comparison of patient’s characteristics

### Postdural puncture headache and initial management

Out of the 63 women who suffered a dural puncture, 36 had PDPH (57.1%). In addition to the headache, 16.7% experienced nausea and vomiting, 19.0% auditory impairment, and 15.9% vision impairment in the immediate postpartum period. An intrathecal catheter was used in 25/63 (39.7%) of those who sustained UDP. An EBP was performed in 15/36 (41.7%) women who exhibited PDPH. At the end of the anesthesia follow-up, 15/36 (41.7%) of women had a complete resolution of their symptoms. The remainder were still experiencing symptoms but improving. The duration of the follow-up was on average 4 days.

### Chronic headache

The risk of chronic headache was higher in the case group when compared to the control group (14.3% versus 4.8%, p=0.049). This finding persisted after adjustment for potential confounders [AOR: 3.67 (1.05, 12.82)] (see table 2). From the nine women presenting chronic headache, four had an EBP. There was no difference in the incidence of chronic headache between those receiving and not receiving EBP (26.7% versus 10.4%, p= 0.2) (see table 3).

**Table 2.** Comparison of outcomes of interest

| Characteristics | Control | Case | p-value* | AOR (95%CI) ‡ |
| --- | --- | --- | --- | --- |
| # Patients | 63 | 63 |  | (Case vs Control) |
| Headache, %(n/N) | 4.8 (3/63) | 14.3 (9/63) | 0.049 | 3.67 (1.05, 12.82) |
| Backache, %(n/N) | 19.1 (12/63) | 39.7 (25/63) | 0.009 | 2.67 (1.25, 5.72) |
| Hearing impairment, %(n/N) | 1.6 (1/63) | 14.3 (9/63) | 0.007 | 9.98 (1.21, 82.62) |
| Visual impairment, %(n/N) | 6.4 (4/63) | 7.9 (5/63) | 0.72 | 1.33 (0.33, 5.40) |
| **Composite outcome, %(n/N)** | 27.0 (17/63) | 52.4 (33/63) | 0.003 | 2.95 (1.42, 6.11) |
|  |  |  |  | Adjust Diff (95%CI) # |
|  |  |  |  | (case vs control) |
| **Total PDI score, median (IQR)** | 4 (0, 17) | 7 (0, 20) | 0.27** | -4.0 (-14.6, 6.62) |

Notes: \* the reported p-values were based on the comparisons between two groups using the probability model with GEE approach to account
for correlation between match pair.
\*\* the p-value was based on the comparison between two groups using quantile regression.
‡ AOR=adjusted Odds ratio (case vs control) based on the conditional logistic regression model to account for the correlation between match pair, adjusted for delivery type.

**Table 3.** Subgroup analysis : Comparison of outcomes of interest in women receiving or not an epidural blood patch

| Characteristics | Non-Epidural blood patch | Epidural blood patch | p-value* | OR (95%CI) ‡<br>(Epidural blood patch vs Non-Epidural blood patch) |
| --- | --- | --- | --- | --- |
| # Patients | 48 | 15 |  |  |
| Headache, %(n/N) | 10.4 (5/48) | 26.7 (4/15) | 0.2 | 3.12 (0.72, 13.63) |
| Backache, %(n/N) | 35.4 (17/48) | 53.3 (8/15) | 0.22 | 2.08 (0.64, 6.74) |
| Hearing impairment, %(n/N) | 12.5 (6/48) | 20.0 (3/15) | 0.43 | 1.75 (0.38, 8.06) |
| Visual impairment, %(n/N) | 8.3 (4/48) | 6.7 (1/15) | 0.99 | 0.79 (0.08, 7.62) |
| **Composite outcome, %(n/N)** | 47.9 (23/48) | 66.7 (10/15) | 0.2 | 2.17 (0.65, 7.32) |
|  |  |  |  | Diff (95%CI) # (case vs control) |
| **Total score, median (IQR)** | 6 (0, 19) | 13.5 (2, 32) | 0.28 | -1.0 (-24.1, 22.1) |

Notes: \* the reported p-values were based on Chi-square or Fisher exact test as appropriate
\*\* the p-value was based on quantile regression.
‡ OR= Odds ratio (case vs control) based on the conditional logistic regression model.

### Chronic lower back pain

The risk of chronic lower back pain was higher in the case group when compared to the control group (39.7% versus 19.1%, p=0.009). The result was consistent after adjustment [AOR: 2.67 (1.25, 5.72)]. There was no difference in the incidence of chronic lower back pain between those receiving and not receiving EBP (53.3% versus 35.4%, p=0.22). Out of the 25 women who had chronic lower back pain, three (12.0%) indicated a probable neuropathic component of their pain. One of those three women received EBP.

### Chronic auditory symptoms

The incidence of chronic auditory symptoms was higher in the dural puncture group compared with the control group [14.3% versus 1.6%, p=0.007; AOR: 9.98 (1.21, 82.62)]. Out of the 9 women in the dural puncture group who reported auditory symptoms, seven self-reported the problem while two had the problem diagnosed by a healthcare professional. The symptom was tinnitus in four of these women and hearing impairment in the five others. The woman in the control group self-reported tinnitus. Women who received EBP and those who did not receive EBP had similar incidence of chronic hearing impairment ((20.0%) versus 12.5%, p= 0.43).

### Chronic visual symptoms

Chronic visual symptoms were reported by (7.9%) women in the control group. Four women had this problem diagnosed by a healthcare professional and one self-reported her symptom. The symptom was decreased visual acuity in 4 women and diplopia in one woman. In comparison, 6.4% of women in the control group had chronic visual symptoms. Two had symptoms that were self-reported and two had their symptoms reported by a healthcare professional. The symptom was decreased visual acuity in all four patients. The difference between the two groups was not significant (p=0.72). There was no difference in visual impairment in those receiving or not EBP (6.7% versus 8.3%, p=0.99). It is worth mentioning that the woman who received the epidural blood patch is the one who suffered chronic diplopia diagnosed by a healthcare professional.

### Composite outcome and Pain Disability Index score

In the control group, 52.4% women developed at least one of the chronic symptoms in comparison to 27.0% women in the control group (p=0.003). The odds of developing at least one of the chronic symptoms in the case group was 195% higher compared to the control group [AOR: 2.95 (1.42, 6.11)]. Women in both groups who developed chronic symptoms completed the Pain Disability Index (PDI) questionnaire to assess the level of disability attributed to those symptoms. The median PDI total score in the case group was similar to the control group [7 versus 4; p=0.27, diff -4.0 (−14.6, 6.62)]. The incidence of women presenting with at least one chronic condition was not different in those receiving or not EBP (66.7% versus 47.9%, p=0.2). Similarly, there was no difference in their median PDI total score [13.5 versus 6, p=0.28; diff -1.0 (−24.1, 22.1)].

## Discussion

Our study suggests that women who sustain UDP during labor are at higher risk of developing chronic headache, chronic lower back pain and chronic auditory symptoms. The use of EBP does not seem to affect the incidence or severity of these outcomes; however, the number of events may be too small to draw that conclusion, and further studies are warranted.

The incidence of PDPH in our cohort (57.1%), as well as the incidence of other acute symptoms accompanying PDPH such as nausea, vomiting, hearing and visual complaints, is in keeping with what has been reported in the literature ^(3)^. The management of PDPH in our cohort may have been more conservative than that reported in similar studies. We performed EBP in 41.7% of women who had PDPH, whereas Binyamin et al ^(25)^, Webb et al ^(5)^ and Ranganathan et al ^(7)^ reported 51.3%, 62.5% and 53.4% respectively.

The incidence of chronic complications in our study is in keeping with those reported in previous similar studies. Webb et al ^(5)^ reported chronic headache in 18% and chronic back pain in 33% of women sustaining UDP. Ranganathan et al ^(7)^ described that 34% of women who sustained UDP developed chronic headache and 58% developed chronic backache, however they did not exclude women with pre-existing symptoms. MacArthur et al ^(16)^ looked at the onset of new headache, migraine or neck pain within three months after childbirth and lasting over six weeks; they reported one or more of these symptoms in 23% of women sustaining UDP in their study. A more recent prospective case-control study by Binyamin et al ^(25)^, where women with previous symptoms were excluded, noted the presence of chronic headache and chronic back pain in 16.1% and 17.9% of women respectively, amongst those who sustained UDP and did not receive EBP.

Similar to what was observed by Ranganathan et al ^(7)^ and Binyamin et al ^(25)^, we did not observe a difference in the incidence of chronic symptoms in women who had an epidural blood patch compared to those who did not. Webb et al ^(5)^, however, reported a lower incidence of both headache and backache in women who received EBP as compared to those who did not. Further studies are warranted to explain these different findings.

Different mechanisms have been proposed to explain the presence of chronic headache following UDP. Chronic cerebral spinal fluid (CSF) leakage has been one of them ^(17)^. Hence, epidural blood patch would be expected to reduce the incidence of this complication ^(18)^. Increased hypersensitivity to substance P ^(19)^, central sensitization ^(20)^ and severe or poorly addressed acute pain resulting in pain memory ^(21)^ are other associated etiologies. The last two mechanisms can also explain the presence of chronic lower back pain. As per our results, the presence of a neuropathic cause seems less likely.

We observed a high incidence of hearing impairment in our cohort, approximately 3 times higher than what was described by Ranganathan et al. ^(7)^ who reported these symptoms in 4.65% of their patients. Hearing loss has been associated with PDPH and spinal anesthesia ^(22)^. It is thought to be secondary to an imbalance of the perilymph in the inner ear ^(23)^. This imbalance would be attributed to the decreased CSF pressure from its leakage.

Diplopia can be a very incapacitating complication of PDPH ^(24)^. Reassuringly, the incidence in our cohort was low, and only one patient developed diplopia. This patient received EBP and it is uncertain if the EBP had any impact on the outcome. In one previous study ^(24)^, the early performance of an EBP showed to prevent progression of visual symptoms if they develop with the PDPH.

We wanted to look at how the chronic headache or lower back pain impacted women’s daily activities since we did not find any studies that looked at this outcome before. We thought it was an important aspect to consider since these women will have to care for their child(ren) in addition to all the other daily activities for many years to come. We were unable to document any significant impact of these complications.

Our study has some limitations. Firstly, we could not control some differences in the acute management of PDPH, given the retrospective nature of our study. Secondly, our study is subject to some respondent bias. We recruited women who sustained UDP between 6 months and 5 years prior to the recruitment; therefore some recall bias from our participants is likely. Another possible bias is the desirability bias whereby women with the most severe symptoms may have been more likely to agree to participate in the study. This could have overestimated the real incidence of chronic headache and backache. Furthermore, we did not have validated questionnaires for the evaluation of chronic visual and chronic auditory disturbance and we had to proceed with a dichotomic “yes” or “no” question to look for them. This could have resulted in a more subjective result for this part of our survey and may have overestimated our results for these two outcomes, which might or not be related to UDP.

## Conclusion

In conclusion, pregnant women who sustain UDP during labour epidural catheter insertion have a higher risk of developing chronic headache, lower back pain and auditory symptoms than those who receive epidural analgesia and do not sustain such complication. The treatment with an EBP does not seem to influence the incidence and severity of these long-term complications, however these results warrant further studies.

## Supporting information

Research Reporting Guideline

## Data Availability

All data produced in the present study are available upon reasonable request to the authors.

## Acknowledgement

Dr. J. Carvalho is supported by the Merit Awards Program from the Department of Anesthesiology and Pain Medicine, University of Toronto

## Notes

**Conflicts of interest** None declared

### Competing Interest Statement

The authors have declared no competing interest.

### Funding Statement

This study was funded by the department only.
No external funding was received.

### Author Declarations

Ethics Review Board at Mount Sinai Hospital in Toronto, Ontario, Canada (REB#20-0278-E; January 20th, 2021) gave ethical approval for this work.

